# Signatures of mast cell activation are associated with severe COVID-19

**DOI:** 10.1101/2021.05.31.21255594

**Authors:** Janessa Tan, Danielle E. Anderson, Abhay P. S. Rathore, Aled O’Neill, Chinmay Kumar Mantri, Wilfried A. A. Saron, Cheryl Lee, Chu Wern Cui, Adrian E. Z. Kang, Randy Foo, Shirin Kalimuddin, Jenny G. Low, Lena Ho, Paul Tambyah, Thomas W. Burke, Christopher W. Woods, Kuan Rong Chan, Jörn Karhausen, Ashley L. St. John

## Abstract

Lung inflammation is a hallmark of Coronavirus disease 2019 (COVID-19) in severely ill patients and the pathophysiology of disease is thought to be immune-mediated. Mast cells (MCs) are polyfunctional immune cells present in the airways, where they respond to certain viruses and allergens, often promoting inflammation. We observed widespread degranulation of MCs during acute and unresolved airway inflammation in SARS-CoV-2-infected mice and non-human primates. In humans, transcriptional changes in patients requiring oxygen supplementation also implicated cells with a MC phenotype. MC activation in humans was confirmed, through detection of the MC-specific protease, chymase, levels of which were significantly correlated with disease severity. These results support the association of MC activation with severe COVID-19, suggesting potential strategies for intervention.

## Introduction

Coronavirus Disease 2019 (COVID-19) is caused by Severe Acute Respiratory Syndrome Coronavirus 2 (SARS-CoV-2), a recently emerged coronavirus that has resulted in an ongoing global pandemic. Clinical disease ranges from asymptomatic to mild to severe, and manifestations include upper respiratory tract symptoms, pneumonia and, in some cases, acute respiratory distress syndrome (ARDS)(1). Fever, cough and anosmia are most commonly experienced at disease presentation and complications involving the vascular system can occur during severe disease(1). The lung is a major target organ of SARS-CoV-2 infection due to abundant expression of the angiotensin converting enzyme 2 (ACE2) receptor, a cellular entry receptor for SARS-CoV-2(2). The virus is typically shed from the nasopharyngeal tract and disseminated by coughing, but it can also be detected in fecal excretions(3). Various mouse and non-human primate (NHP) models have been utilized to study COVID-19(4). Non-human primates (NHPs) and human ACE2 (hACE2) knock-in mice both have been shown to experience infection and recapitulate human signs of disease in the lung, including lung pathology(4). In human autopsy studies of severe disease, infiltration of mononuclear cells in the lung tissue concurrent with edema and hemorrhage are frequently described(5). It is believed that lung pathology during COVID-19 is immune-mediated and compounded by the infiltration of monocytes, neutrophils and subsets of T cells(6). Interestingly, perturbations in the numbers of granulocytes in the blood, such as neutrophils and eosinophils have also been shown to be associated with severe disease(1, 7, 8).

Another granulocyte that responds to viral infections and is found in the lung tissue is the mast cell (MC). MCs are long-lived granulated immune cells that are present in both connective and mucosal tissues(9). In adults, MCs are thought to be derived from precursor cells circulating in the blood, known as MC progenitors, but they are only found in mature form in tissues(10), making them difficult to study in humans. Tissue resident MCs have a mature phenotype and express a variety of pathogen recognition molecules on their surface and inside cytosolic compartments(11, 12). Their granules are loaded with pre-formed mediators such as histamine, serotonin and unique MC-specific proteases, chymase and tryptase, among others. Some of their mediators, including soluble cytokines and lipid mediators, may be also produced by other granulocytes and immune cells(13). MC-derived products not only promote tissue inflammation through the recruitment of cells such as monocytes, neutrophils and T cells, they also have significant effects on vascular permeability and vasomotor control(9, 12, 14). The influence of MCs on vasomotor control, including vasoconstriction and vasodilation, may also contribute to hypoxia that occurs through shunting, which can influence vascular and tissue integrity(15). The tissue-specific microenvironment where a MC resides influences its phenotype. For example, MCs in the atopic lung express higher levels of IgE receptor, FcεR1 than in the skin(16, 17) and lung MCs are well characterized to contribute to pathological lung inflammation during conditions such as asthma(18). MCs are known to coordinate effective immune responses against invading pathogens, including viruses(12) but their activation has also been linked to severe tissue damage, such as during dengue virus (DENV) infection(19). In the lung, MC hyperplasia has also been reported during respiratory syncytial virus or parainfluenza virus infections(20, 21) and therapeutic stabilization of MCs was shown to reduce lung lesions in a model of highly pathogenic H5N1 influenza infection(22). However, sustained and systemic activation of MCs could also result in severe pathologies such as coagulation disorders and vascular leak. For example, MC-specific products such as chymase have been shown to be predictive of dengue hemorrhagic fever and the severity of vascular leakage and coagulopathy that characterize severe disease (19, 23, 24). MCs are present both in the nasal mucosae as well as in the deeper lung tissue where SARS-CoV-2 infection occurs; however, it is unknown whether MCs respond to highly pathogenic coronaviruses or if they could be involved in exacerbating the severe inflammation seen in SARS-CoV-2 infection.

In this study we aimed to assess MC activation in response to SARS-CoV-2 infection. Using mouse and NHP models of COVID-19 we identified wide-spread MC degranulation in both acute and convalescent lung tissues. In a human cohort, prospective analysis of the transcriptional signatures of MC-precursors were highly enriched in the blood of patients who presented with severe COVID-19 disease, suggesting modulation of this cell type during disease, as were several host response pathways for prominent MC-derived products. Furthermore, the MC-specific product, chymase was significantly elevated in the sera of SARS-CoV-2 infected patients confirming human MC activation during COVID-19 and supporting the likelihood that MCs contribute to severe COVID-19 disease.

## Results and Discussion

### MC degranulation coincides with lung pathology in animal models of COVID-19

Given the association of MCs with chronic airway inflammation, their immune sentinel role for certain viral pathogens, and knowing that severe lung inflammation also characterizes COVID-19, we first questioned whether MCs are activated in animal models of SARS-CoV-2 infection. We used an established mouse model where the receptor for SARS-CoV-2, hACE2, is delivered to the lungs using an adenovirus vector(25). After hACE2-AAV inoculation, mice were infected with SARS-CoV-2 (**Fig. 1A**). Blood was collected at multiple time points to assess MC-associated inflammatory products and tissues were collected on days 5 and 7 post-infection for virus quantification by PCR. Mice showed the highest infection burden in the lungs, but for at least some of the animals, SARS-CoV-2 could also be detected in the spleen, liver, kidney, brain, and bone marrow (**Fig. 1B**), while the brachial lymph nodes were PCR-negative at both time points (**Fig. S1**). Tissue histology revealed degranulation of MCs in the airways, as shown in a representative image of the trachea at day 5 post-infection (**Fig. 1C**), where toluidine blue staining of MC granules indicated extensive degranulation that coincided with edema in the tissue. In contrast, granulated resting MCs can be observed in control trachea tissue (**Fig. 1C**). The trachea tissue from control uninfected animals also appears healthy and compact, while the thickness of the trachea tissue in SARS-CoV-2 infected animals appears increased as a result of inflammation and swelling (**Fig. 1C**). To provide a quantitation of MC activation, we also measured serum levels of the mouse chymase MCPT1, which is a MC-specific protease that can be used as a biomarker of MC activation(19). MCPT1 levels were significantly elevated days 1, 3 and 5 post-SARS-CoV-2 infection and remained high, but trending lower on day 7 (**Fig. 1D**), which we noted also coincided with reductions in viral burden in the tissues (**Fig. 1B**). The evidence of MC degranulation in the airways combined with systemically elevated MC products indicates that SARS-CoV-2 induces substantial activation of MCs during infection in vivo.

**Figure 1.**
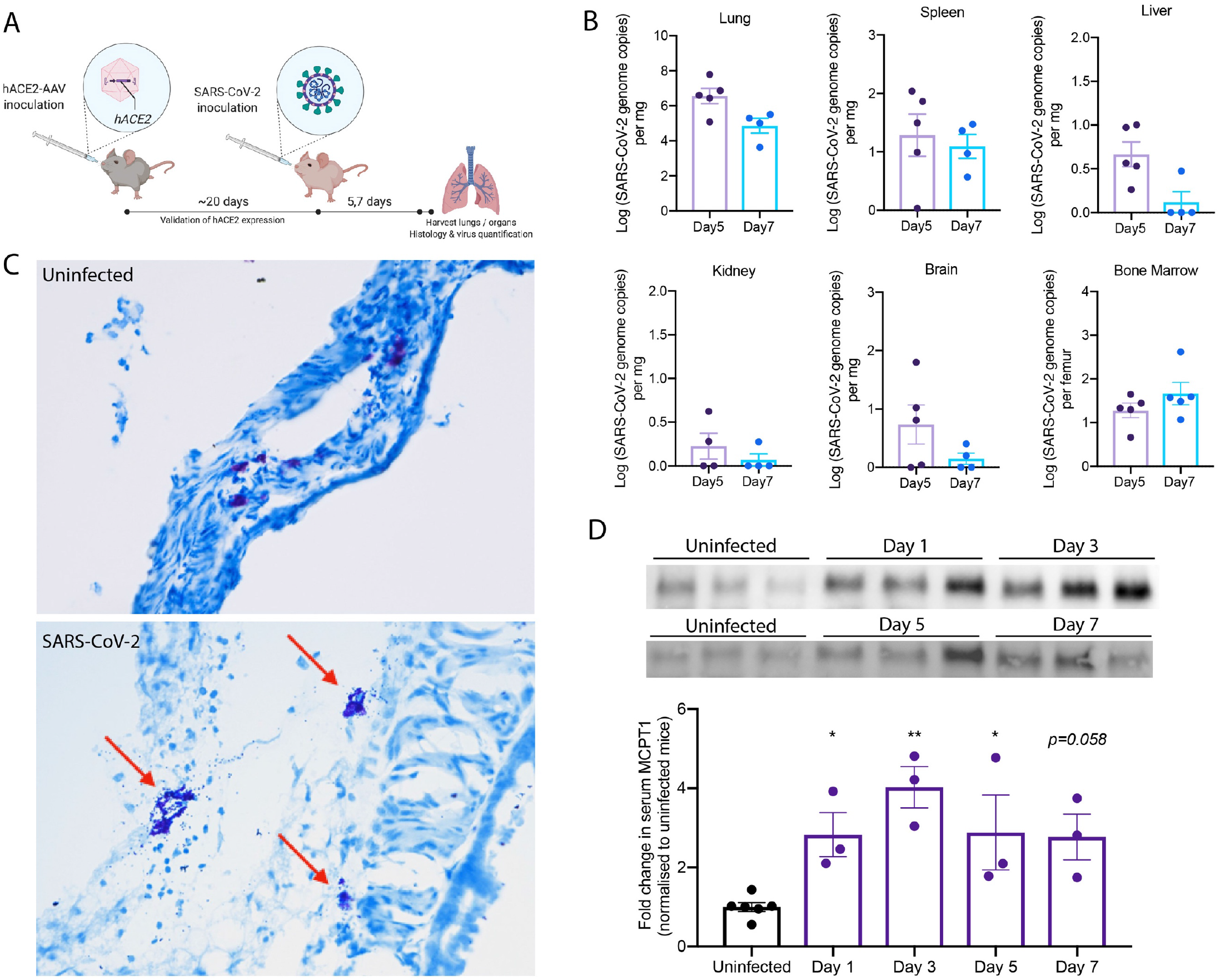
Degranulation of MCs in SARS-CoV-2 infected mice. (**A**) C57BL/6 mice were inoculated intranasally with *hACE2*-AAV to induce *hACE2* expression in the airways. SARS-CoV-2 (2×10^7^ TCID_50_) TCID-50) was inoculated intranasally into *hACE2*-AAV C57BL/6 mice. Blood was taken on days 1, 3, 5, and 7, and organs were harvested after 5 or 7 days for histology and virus quantification. (**B**) Virus quantification from the organs harvested shows detection in the lung, spleen, liver, kidney, brain, and bone marrow both Days 5 and 7. (**C**) Histology images of toluidine blue-stained trachea sections from uninfected and SARS-CoV-2 infected mice. Degranulating MCs (red arrow) could be observed in SARS-CoV-2 infected mice as well as tissue edema and airway narrowing. (**D**) Western blot images after chymase detection in serum Days 3, 5 and 7 post-infection shows systemic elevation of chymase, which was quantitated by densitometry from 3 individual mouse samples and presented as fold-increase over uninfected controls. Error bars represent the SEM. Chymase was significantly elevated in serum of infected mice compared to uninfected controls, determined by 1-way ANOVA with Dunnett’s post-test; ^*^*p*<0.05, ^**^*p*<0.01. Non-significant *p*-values below p=0.01 are shown on the graph.

We next aimed to validate the MC activation phenotype in the non-human primate (NHP) model, which is thought to more closely replicate the signs and symptoms of human SARS-CoV-2 infection(4). For this, cynomolgus macaques were infected with 3×10^6^ TCID-50 of SARS-CoV-2 virus intra-tracheally and they were monitored with minimal interventions for 21 days prior to necropsy (**Fig. 2A**). Throughout the study the animals were generally active, alert, and responsive. There were no significant changes in body weight or temperature during the study (**Fig. S2**). Two NHPs (#6699 and 6727) displayed appetite loss, and one was given subcutaneous fluids. SARS-CoV-2 could be detected in the nasal rinse or swab of all NHPs at multiple time points during acute infection, as well as in the throat swab and lung lavage at least one time point post-infection (**Fig. 2B**). Additionally, 3 of 4 NHPs were positive by rectal swab and 1 also had detectable SARS-CoV-2 by eye swab (**Fig. 2B**). In support of active infection, all NHPs seroconverted by day 14 (**Table S1**). At the time of necropsy on day 21, evidence of severe lung disease was apparent, with all displaying damage to the lung tissue including areas of hemorrhaging visible on the lungs and fluid accumulation in the lungs (**Fig. 2C-D**). Additionally, one NHP had blood clots inside the lungs and 50% of NHPs had areas of black necrotic patches on the lungs (**Fig. 2C-D**), indicating severe virus-induced pathology. RNA was extracted from lung tissue from each NHP and all samples were PCR-negative for SARS-CoV-2. Interestingly, upon necropsy, NHP #6727 had detectible virus in the cerebrospinal fluid (CSF). These findings suggested that the NHPs in this study experienced ongoing inflammation and tissue damage even after the resolution of active infection.

**Figure 2.**
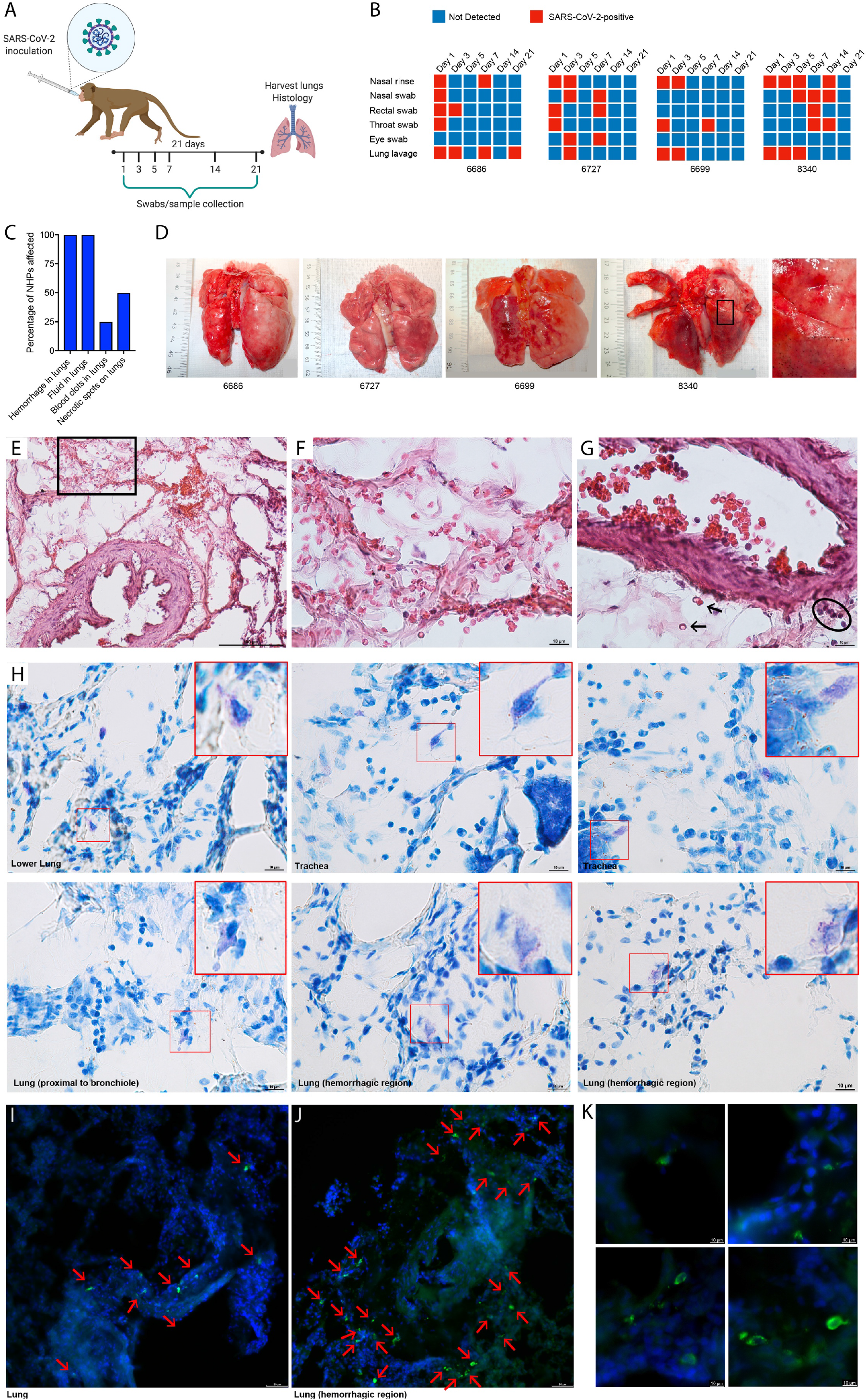
Widespread activation of MCs coinciding with lung pathology in NHPs. (**A**) Cynomolgus macaques were infected intratracheally with SARS-CoV-2 and monitored for 21 days prior to necropsy. (**B**) Viral detection was determined by PCR at regular intervals post-infection in swabs from multiple mucosal tissues, lung lavage, and nasal rinses. All NHPs were positive for SARS-CoV-2 infection multiple days after inoculation. (**C**) Abnormal findings related to lung tissue observed at the time of necropsy were recorded and effected all animals. (**D**) Images of NHP lungs at the time of necropsy show areas of hemorrhaging and necrotic spots on the lung surface. Boxed region is enlarged. (**E)** Histological assessment of lung tissues by H&E staining shows hemorrhaging of the tissue and free RBCs within the lung alveolar spaces. (**F**) Inset corresponding to the boxed region of panel H. (**G**) Some RBCs in the tissue proximal to a blood vessel are indicated by arrows and cellular infiltrates are circled. (**H**) Multiple examples of degranulating or hypogranulated MCs are provided, observed in toluidine blue stained lung tissue sections. The MCs are enlarged in the red-outlined insets. For (**I-K**), lung sections were stained for MC heparin to indicate the location of MC granules (green) and DAPI to identify cellular nuclei and tissue structures. MCs are indicated with red arrows. (**I**) MCs were observed degranulating in the lung of SARS-CoV-2 infected primates in sections of a biopsy of lung tissue that did not have overt hemorrhaging visible on the lung surface at necropsy. (**J**) MCs appear more densely packed in the lung biopsy from a hemorrhagic lobe of the lung and again, degranulation is observed based on staining for MC-heparin. (**K**) Images of degranulating MCs are presented at higher magnification.

Histological assessments of lung tissue showed severe damage to the airways and lung-associated vasculature that coincided with activation of MCs in tissues. Signs of hemorrhage were present in the lung tissue, where red blood cells (RBCs) were observed in the extravascular space, both trapped within the alveoli, which were occasionally abnormally thickened (**Fig. 2E-F, S3**), as well as near blood vessels (**Fig. 2G, S3**). Proximal to blood vessels there was also evidence of infiltration of immune cells into the tissue (**Fig. 2G**) and fibrin deposition (**Fig. S3**). In multiple locations within the lung, including in the trachea and the lower lung lobes, as well as near bronchi and near alveolar spaces, hypodense MCs could be observed after staining of tissue sections with toluidine blue, suggesting their recent degranulation (**Fig. 2H**). Free granules could be observed extracellularly near MCs (**Fig. 2H**), also indicating degranulation. This widespread activation of MCs was confirmed by fluorescence staining to detect heparin-containing granules in the lung tissue (**Fig. 2I-K**). We noted that activated MCs were especially densely located and degranulating within the hemorrhagic regions of the infected lung tissue (**Fig. 2H, J**). At higher magnification, free granules could be observed near hypogranulated MCs (**Fig. 2K**), also indicating recent degranulation. These results support that SARS-CoV-2 infected NHPs experience lung pathology involving hemorrhagic manifestations and widespread MC activation, which persists to late time points in the disease course.

### Signatures of MC transcriptional activation are associated with severe COVID-19

In healthy humans, MC precursors make up a minor component of the blood, ∼0.005% of cells(26). MCs are known to have a unique transcriptional profile that clusters separately from other immune cells and gene expression patterns have been identified that are either MC-specific or that typify both MCs and basophils(27). Although MCs are not present in mature form in the blood, we considered that their activation in peripheral tissues could influence the MC precursors or lead to transcriptional activation profiles in immune cells that are consistent with responses to systemically elevated MC-associated products. To investigate this, we examined whole blood transcriptomics data from a cohort of 4 mild and 6 severe COVID-19 patients, where clusters of genes that were temporally modulated during severe disease progression and resolution were identified (28). Consenting patients were prospectively recruited and were defined as severe on the basis of requiring supplemental oxygen during hospitalization. In the patients with severe disease, the gene expression levels were monitored from -4 days to 13 days, relative to the day when their condition peaked in severity of respiratory distress, which was defined as time=0 (28). Interestingly, many genes associated with the MC lineage (**Fig. 3A-B**) or MC and also basophil lineages (**Fig. 3C-D**) were differentially modulated in the blood of human COVID-19 patients with severe disease (p-value < 0.05; q-value < 0.05; likelihood ratio test). Upregulation of several genes associated with the MC- or MC/basophil transcriptional signature(27) occurred during the acute phase of severe disease (**Fig. 3A,C**), while others were differentially regulated at the time of disease resolution (**Fig. 3B,D**). The increased MC gene expression changes that were observed during the acute phase of disease tracked tightly with respiratory function and resolved commensurate with respiratory improvement **(Fig. 3E)**. In contrast, these MC-associated transcripts were not collectively changed temporally throughout the period of monitoring in mild COVID-19 presentation (**Fig S4A-E**), although some genes that were associated with these signatures were still modulated, but to a lesser extent than in severe patients (**Fig. S4A-D**). Pathway analysis of the temporally modulated genes over the disease course of severely ill patients revealed significant perturbation of pathways downstream of key MC-associated immune receptors (**Fig. 3F**) such as KIT (**Fig. 3G**), the receptor for stem cell factor, which is an important stem cell-associated gene that is retained on MC precursors and mature MCs and regulates MC survival and proliferation(29), and FcεRI (**Fig. 3H**), which is upregulated with MC maturation, although also expressed by other cell types such as basophils(26, 27). These data show an enrichment of MC-associated transcripts in patients with severe COVID-19 and support a potential role of MCs in shaping disease severity.

**Figure 3:**
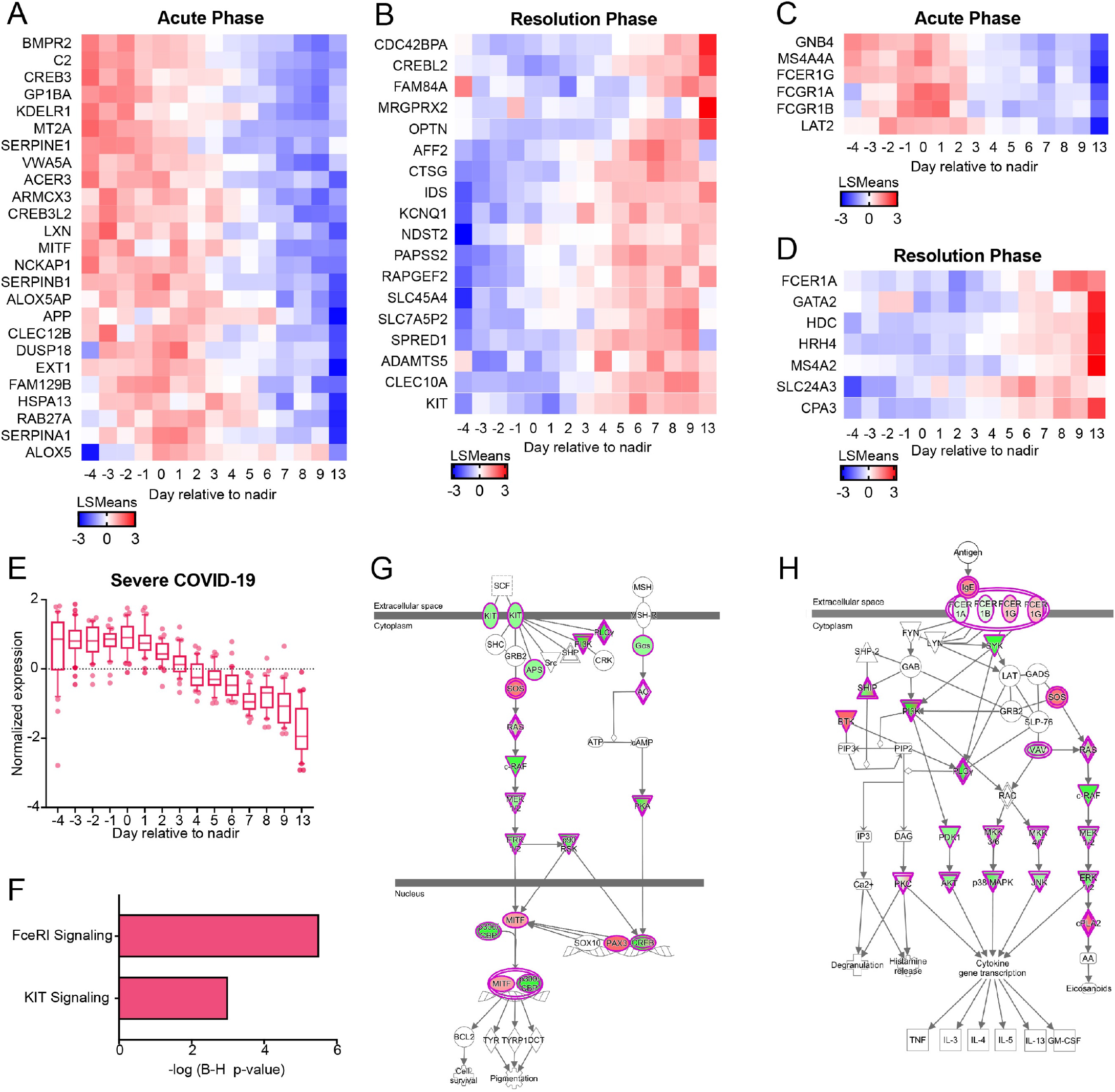
Transcriptional signatures of MC associated genes with severe COVID-19. Genes associated with a (**A-B**) MC-specific or (**C-D**) MC/basophil phenotype that were significantly regulated in PBMCs of severe COVID-19 patients. Heatmap shows the LSmean expression values of MC-specific or MC/basophil phenotype genes in severe COVID-19 patients (n=6) at the various days relative to the peak severity with respect to respiratory function (day 0). Clusters of genes that were significantly upregulated during the acute phase (**A, C**) or resolution phase (**B, D**) are presented. (**E**) Normalized expression levels of the MC-specific genes shown in A and C over time, in the severe COVID-19 patients. (**F**) Pathway analysis indicates a significant perturbation of pathways associated with MC function and/or MC-precursor maturation. Gene network analysis for the significantly modulated pathways (**G**) KIT and (**H**) FcεRI are shown. Red indicates the genes with increased expression during the acute phase, whereas green indicate genes with increased expression during the resolution phase.

### Confirmation of MC activation in human COVID-19 patients

We next examined mild and severe COVID-19 patient blood for evidence that could indicate responses to MC products and for biomarkers of MC activation. We noted that in addition to the significant modulation of pathways associated with MC identity and maturation (**Fig. 3F-H**), pathway analysis of the whole blood transcriptomics from severe patients also revealed significant modulation of pathways associated with responses to well-established MC products (**Fig. 4A**). For example, Gap and adherens junction signaling, which are influenced by MC proteases to promote vascular permeability(14), were activated, as was signaling downstream of important, albeit not cell-specific, MC products, such as VEGF, TNF, Endothelin 1, and Eicosanoids (**Fig. 4A**). We also noted a significant influence on the renin-angiotensin pathway, which is intriguing since chymase mediates angiotensin-converting enzyme (ACE)-independent angiotensin II production (**Fig. 4A**).

**Figure 4:**
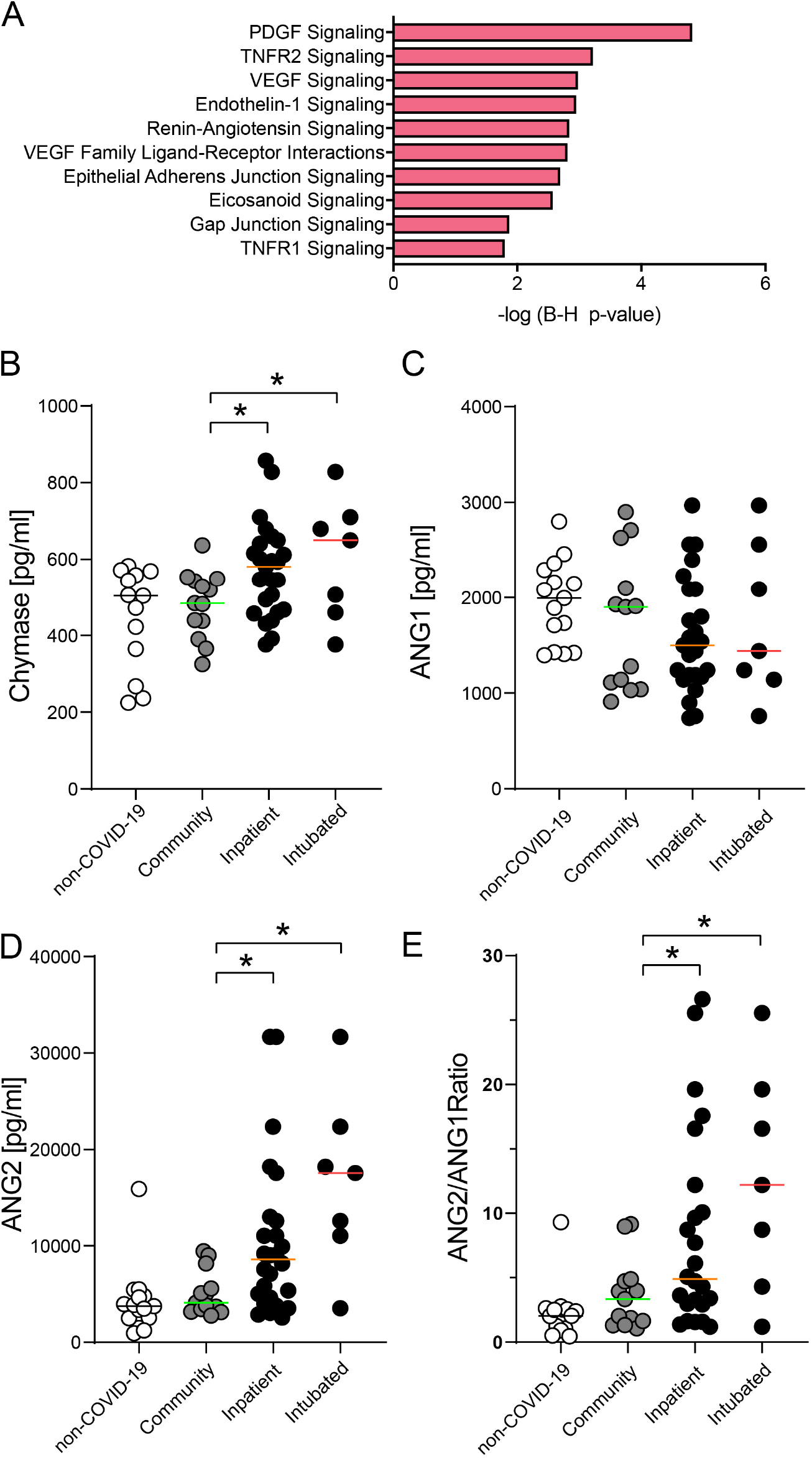
MC products and activation pathways associated with severe COVID-19. (**A**) Pathway analysis indicates a significant perturbation of pathways associated with host-responses to characteristic MC products. (**B**) Plasma chymase levels are increased in hospitalized patients “inpatient” compared to mild “community” patients and in intubated patients compared to mild patients. (**C**) No significant changes in ANG1 levels in inpatient and intubated patients compared to community cases and non-COVID-19 controls. (**D**) Significantly increased plasma ANG2 levels and (**E**) ANG2/ANG1 ratios in inpatients compared to community patients. One-way ANOVA was performed and considered significant for post-test p-values <0.05. For panels, **B-E**, non-COVID-19 patients n=12-15; For community, n=13; for inpatients n=24; n=7.

To confirm the activation of MCs in humans, we also measured plasma chymase levels in two other separate cohorts of COVID-19 patients. In the first cohort, the levels of plasma chymase were measured in 13 patients that presented with mild disease and were treated as outpatients, (“community”), and compared to the plasma chymase levels of 24 patients, in whom severe disease necessitated hospitalization (“inpatients”), or to non-COVID-19 controls (**Fig. 4B**). The WHO 10-point median clinical disease severity(30) in inpatients was 6 (25^th^ and 75^th^ interquartile, 5 and 7.25), including 7 intubated patients and 3 patients with lethal outcomes. As non-COVID-19 controls we obtained baseline plasma samples (after the induction of anesthesia, before incision) from patients who underwent coronary bypass surgery. This cohort was chosen since they have many of the risk factors of COVID-19 patients and are of a similar age. In both mild and severe COVID-19 cases, the plasma sample was collected at the time of diagnosis for the majority of patients. These results indicated that hospitalized COVID-19 inpatients have significantly higher levels of plasma chymase compared to community cases (**Fig. 4B**). This difference was emphasized when the chymase levels in community cases were compared to hospitalized patients that required intubation (**Fig. 4B**). Published reports highlight the importance of microvascular abnormalities in defining COVID-19 severity(31) and are supported by the demonstration of alveolar edema and hemorrhagic lesions in our murine and NHP models. MC activation has direct impact on vascular function and integrity and, therefore, we tested if MC activation was linked to vascular barrier dysfunction. For this, we measured Angiopoietin (Ang)-1 and -2 levels as markers of endothelial activation, which are strongly linked with disease severity in ARDS(32) and COVID-19(32) and found no change in Ang1 levels (**Fig. 4C**), yet higher Ang2 levels (**Fig. 4D**), resulting in higher Ang2/Ang1 ratios (**Fig 4E**) in hospitalized and intubated vs. community COVID-19 cases. We also recruited a smaller number of COVID-19 patients in Singapore. Indeed, this second cohort of COVID-19 patients also had elevated chymase that was much higher than healthy controls and also averaged higher than the concentrations detected in acute dengue patients (**Fig. S5**). These data were not stratified by severity due to the smaller cohort size but support the activation of MCs in human COVID-19 patients. Taken together, human chymase detection confirms that elevated chymase and heightened MC activation are associated with severe COVID-19.

Our results indicate that MCs are strongly activated by SARS-CoV-2 infection in vivo in animal models, and that their levels of activation are significantly associated with severe COVID-19 disease in humans. The activation response involves a degranulation and release of MC-associated pre-formed mediators, which was confirmed visually by imaging of tissue sections as well as quantitatively by detection of MC-specific chymase in the serum. MCs are present in the lung tissue, even prior to birth, and they are important for regulating lung tissue inflammation during homeostasis and disease(33). The heightened levels of persistent activation of MCs that we detected through the acute phase of natural and experimental SARS-CoV-2 infections are likely to be important for amplifying inflammation, which could be detrimental to recovery from infection and return to tissue homeostasis following infection clearance. In mouse and NHP lung tissue, MCs were observed to be strongly degranulating, and their increased density and morphological appearance of activation was associated with areas of tissue damage characterized by edema, hemorrhaging and necrosis. This is consistent with the role for MCs in mediating inflammation and pathology in the lung that was suggested by mouse models of highly pathogenic influenza(22, 34). The persistent activated state observed in the lungs of NHPs, with sustained evidence of MC activation at the relatively late time point post-infection when samples were no longer PCR-positive for virus detection, suggests that ongoing inflammation in the tissue may occur even after infection has resolved systemically. The late phase activation of MCs subsequent to infection clearance and establishment of a humoral adaptive immune response could suggest a role for MCs in the sustained inflammatory response that limits disease resolution. Consistent with this, all NHPs seroconverted by 3 weeks post-SARS-CoV-2 infection and beginning as early as 1 week post-infection. Interestingly, IgGs targeting various self-antigens including IFNs, phospholipids and cytokines, as well as heightened total IgE levels have been detected in severe COVID-19 patients(35-38). MCs respond to antibodies in unique ways when triggered by antigen/antibody immune complexes. Classically, known for its activation by crosslinking of IgE-FcεRI in the presence of an antigen, MCs can also be activated by IgG immune complexes owing to their surface expression of activating FcγRs(39-41). In addition to genes that are consistent with a MC-specific transcriptional profile, we also found significant upregulation of pathways typifying both MCs and basophils (27) in severe compared to mild COVID-19 patients, such as multiple Fc receptors. Whether MCs can also be activated by autoantibodies that are evoked in the absence of an active infection remains to be elucidated and might be relevant to long COVID-19 with persistent symptoms.

We also observed transcriptional responses of severe COVID-19 patients that coincide with peak respiratory distress and which point to the enhanced function or abundance of cells having a MC-like phenotype. Pathways characteristic of pro-inflammatory responses of responder cells to MC-derived products were also modulated. Since mature MCs are not present in the blood(10) and only present in tissues, the MC associated phenotype described here is likely more consistent with MC precursors than with mature MCs. We noted that transcripts of proteins that are specific to mature granulated MCs, such as chymase, were not identified as a component of the MC-associated transcriptional profile that was induced along the time course of peak severity. These results are highly suggestive of the expansion or increased maturation of MC precursors in the blood, but further studies are needed to fully understand the responses of this cellular compartment to infection. A limitation here is the potential to only monitor transcriptional responses in the human blood, yet, our animal model data supports that there is expansion of MCs in the lung tissue as well. This is consistent with the observation in a murine model of H1N1 influenza infection where recruitment and maturation of MC progenitors in the lung was suggested to occur approximately 2 weeks after the infection(34). We also noted an unusually high density of MCs in damaged and hemorrhagic regions NHP lungs at 3 weeks post SARS-CoV-2 infection. Increased transcriptional upregulation of the chemokine CXCR2 in the blood of severe COVID human patients is also suggestive of MC precursor migration into lung, as was seen in the context of other diseases(42).

Similarly, other studies have identified transcriptional signatures of granulocyte activation as well as increases in cells such as neutrophils, eosinophils and basophils and T cells in the blood or lung tissue itself in severe COVID-19(1, 6-8). As tissue-resident cells, MCs are considered sentinels and they can promote the trafficking of many of these cell types into tissues during both allergic and infection-induced inflammation(9, 12, 43).

Aside from the lung-associated pathologies of COVID-19, some individuals also experience other hematological changes and cardiovascular events, including intra-vascular coagulation, endothelial damage with ischemic complications, the development of rashes that could be accentuated by damaged microvasculature, and increased incidence of myocardial infarction(1, 44). These effects on the vasculature and cardiovascular system are also consistent with the effects of MCs in other sterile inflammatory conditions. MCs line the blood vessels within tissues(14), which not only places them in a location where they can directly exert their effects on the vasculature, but also where their mediators can gain access to the blood. We observe that lung SARS-CoV-2 inoculation in mice and humans both results in increased levels of MC-specific chymase, on a systemic level. In the renin-angiotensin system, MC-chymase is a potent converter of angiotensin I to angiotensin II, which regulates microvascular blood flow and systemic blood pressure(45-47). However, production of chymase by MCs is also associated with vascular diseases. For example, in atherosclerotic aorta, angiotensin II activity was largely ACE-independent and dependent on chymase(48) and increased expression of chymase in the lung was associated with early pulmonary vascular disease(49). Notably, higher levels of angiotensin II in the plasma of COVID-19 patients are correlated with lung injury suggesting its involvement in the tissue damage(50). Moreover, angiotensin II could increase the expression of endothelial-specific receptor tyrosine kinase (TIE2) ligand, Ang2(51). An imbalance of Ang2/1 is known to be associated with vascular leakage and coagulation in other diseases(52). We observed increased plasma levels of Ang2 and increased ratio of Ang2/Ang1 in severe COVID-19 patients compared to milder COVID-19 patients or healthy controls. Interestingly, we also observed transcriptional responses of the angiotensin pathway were substantially perturbed in the peripheral blood of severe COVID-19 patients. As exemplified by the endothelial activation in severe COVID-19, this highlights a potential causal role of MC activation in critical features of COVID-19 disease, including abnormalities of pulmonary blood flow leading to shunting and hypoxemia or loss of endothelial integrity leading to tissue edema. Notwithstanding its role as an angiotensin converting enzyme, a more direct effect of chymase in cleaving endothelial tight junctions or potential contributions of other MC products, such as tryptase and serotonin(12), in COVID-19 related vascular pathologies cannot be ruled out. As such, we found intriguing parallels to DENV as another virus that induces MC activation. Although DENV does not specifically infect the lung, DENV infection is also characterized by increases in microvascular permeability and bleeding, which are augmented through the actions of MCs. It is noteworthy that in dengue, MCs play an important role in limiting virus burden in early disease, but drive clinical deterioration in disseminated disease(12). As a result, drugs targeting MCs and their products are promising as a therapeutic strategy to prevent severe clinical courses in DENV infection and may bear similar promise in preventing severe COVID-19, which warrants further evaluation.

## Methods

### Study approvals

All mouse and primate studies were approved by the SingHealth Institutional Animal Care and Use Committee of the SingHealth Experimental Medicine Centre (SEMC). The data associated with human transcriptional responses was approved by the SingHealth Combined Institutional Review Board (CIRB 2017/2374). COVID-19 patients were recruited at Duke University in accordance with protocols reviewed and approved by the Duke University Health System IRB (Pro00100241) while human COVID-19 patient studies in Singapore were approved by the Domain Specific Review Board, Domain E for National University Hospital (#2020/00120) and the National University of Singapore IRB (NUS-IRB-2021-186).

Additional Supplemental Methods accompany the study.

## Data Availability

All data needed to evaluate the conclusions are included in the manuscript and additional data maybe requested.

https://www.ebi.ac.uk/arrayexpress/experiments/E-MTAB-9721/

## Authors contributions

JT, DA, APSR, AO, CKM, and WAAS performed experiments and data analyses. Data were interpreted by JT, DA, APSR, KRC, JK and ALS. ALS and APSR wrote the manuscript with contributions by JT. The primate study was designed, funded and conducted by DA with contributions from AEZK and RF. Histological assessments in mice and primates were done by APSR. hACE2-AAV mouse model was established and validated by CL, CWC and LH. Transcriptional analysis was performed by KRC with additional interpretation by ALS. Human clinical sample collection and patient assessments were performed by JL, SK, PT, TB and CW. Funding for the study was obtained by LH, JK, and ALS. The project was conceived and supervised by ALS. All authors reviewed the manuscript and provided feedback on it.

## Data and materials availability

All data needed to evaluate the conclusions in the paper are present in the paper and/or the Supplementary Materials. Additional data related to this paper may be requested from the authors.

## References

1. St John AL & Rathore APS (2020) Early Insights into Immune Responses during COVID-19. J Immunol.

2. Hoffmann M, et al. (2020) SARS-CoV-2 Cell Entry Depends on ACE2 and TMPRSS2 and Is Blocked by a Clinically Proven Protease Inhibitor. Cell.

3. Patel KP, et al. (2020) Transmission of SARS-CoV-2: an update of current literature. Eur J Clin Microbiol Infect Dis 39(11):2005–2011.

4. Munoz-Fontela C, et al. (2020) Animal models for COVID-19. Nature 586(7830):509–515.

5. Xu Z, et al. (2020) Pathological findings of COVID-19 associated with acute respiratory distress syndrome. Lancet Respir Med 8(4):420–422.

6. Song JW, et al. (2020) Immunological and inflammatory profiles in mild and severe cases of COVID-19. Nat Commun 11(1):3410.

7. Zhang JJ, et al. (2020) Clinical characteristics of 140 patients infected with SARS-CoV-2 in Wuhan, China. Allergy 75(7):1730–1741.

8. Vitte J, et al. (2020) A Granulocytic Signature Identifies COVID-19 and Its Severity. J Infect Dis 222(12):1985–1996.

9. Abraham SN & St John AL (2010) Mast cell-orchestrated immunity to pathogens. Nat Rev Immunol 10(6):440–452.

10. Ribatti D & Crivellato E (2014) Mast cell ontogeny: an historical overview. Immunol Lett 159(1-2):11–14.

11. St John AL & Abraham SN (2013) Innate immunity and its regulation by mast cells. J Immunol 190(9):4458–4463.

12. Rathore AP & St John AL (2020) Protective and pathogenic roles for mast cells during viral infections. Curr Opin Immunol 66:74–81.

13. Wernersson S & Pejler G (2014) Mast cell secretory granules: armed for battle. Nat Rev Immunol 14(7):478–494.

14. Kunder CA, St John AL, & Abraham SN (2011) Mast cell modulation of the vascular and lymphatic endothelium. Blood.

15. Nielsen HB (2003) Arterial desaturation during exercise in man: implication for O2 uptake and work capacity. Scand J Med Sci Sports 13(6):339–358.

16. Andersson CK, et al. (2011) Alveolar mast cells shift to an FcepsilonRI-expressing phenotype in mild atopic asthma: a novel feature in allergic asthma pathology. Allergy 66(12):1590–1597.

17. Fujisawa D, et al. (2014) Expression of Mas-related gene X2 on mast cells is upregulated in the skin of patients with severe chronic urticaria. J Allergy Clin Immunol 134(3):622–633 e629.

18. Brown JM, Wilson TM, & Metcalfe DD (2008) The mast cell and allergic diseases: role in pathogenesis and implications for therapy. Clin Exp Allergy 38(1):4–18.

19. St John AL, Rathore AP, Raghavan B, Ng ML, & Abraham SN (2013) Contributions of mast cells and vasoactive products, leukotrienes and chymase, to dengue virus-induced vascular leakage. eLife 2:e00481.

20. Sorden SD & Castleman WL (1995) Virus-induced increases in bronchiolar mast cells in Brown Norway rats are associated with both local mast cell proliferation and increases in blood mast cell precursors. Lab Invest 73(2):197–204.

21. Castleman WL, Sorkness RL, Lemanske RF, Jr., & McAllister PK (1990) Viral bronchiolitis during early life induces increased numbers of bronchiolar mast cells and airway hyperresponsiveness. Am J Pathol 137(4):821–831.

22. Han D, et al. (2016) The therapeutic effects of sodium cromoglycate against influenza A virus H5N1 in mice. Influenza Other Respir Viruses 10(1):57–66.

23. St John AL & Rathore APS (2019) Adaptive immune responses to primary and secondary dengue virus infections. Nat Rev Immunol 19(4):218–230.

24. Tissera H, et al. (2017) Chymase is a Predictive Biomarker of Dengue Hemorrhagic Fever in Pediatric and Adult Patients. J Infect Dis.

25. Israelow B, et al. (2020) Mouse model of SARS-CoV-2 reveals inflammatory role of type I interferon signaling. J Exp Med 217(12).

26. Dahlin JS, et al. (2016) Lin-CD34hi CD117int/hi FcepsilonRI+ cells in human blood constitute a rare population of mast cell progenitors. Blood 127(4):383–391.

27. Dwyer DF, Barrett NA, Austen KF, & Immunological Genome Project C (2016) Expression profiling of constitutive mast cells reveals a unique identity within the immune system. Nat Immunol 17(7):878–887.

28. Ong EZ, Kalimuddin, S., Chia, W.C., Ooi, S.H., Koh, C.W.T., Tan, H.C., Zhang, S.L., Low, J.G., Ooi, E.E., Chan, K.R. (in press) Temporal dynamics of the host molecular responses underlying severe COVID-19 progression and disease resolution. Ebiomedicine.

29. Galli SJ, Tsai M, & Wershil BK (1993) The c-kit receptor, stem cell factor, and mast cells. What each is teaching us about the others. Am J Pathol 142(4):965–974.

30. Characterisation WHOWGotC & Management of C-i (2020) A minimal common outcome measure set for COVID-19 clinical research. Lancet Infect Dis 20(8):e192–e197.

31. Ackermann M, et al. (2020) Pulmonary Vascular Endothelialitis, Thrombosis, and Angiogenesis in Covid-19. N Engl J Med 383(2):120–128.

32. Gutbier B, et al. (2018) Prognostic and Pathogenic Role of Angiopoietin-1 and -2 in Pneumonia. Am J Respir Crit Care Med 198(2):220–231.

33. Msallam R, et al. (2020) Fetal mast cells mediate postnatal allergic responses dependent on maternal IgE. Science 370(6519):941–950.

34. Zarnegar B, et al. (2017) Influenza Infection in Mice Induces Accumulation of Lung Mast Cells through the Recruitment and Maturation of Mast Cell Progenitors. Front Immunol 8:310.

35. Lucas C, et al. (2020) Longitudinal analyses reveal immunological misfiring in severe COVID-19. Nature 584(7821):463–469.

36. Bastard P, et al. (2020) Autoantibodies against type I IFNs in patients with life-threatening COVID-19. Science 370(6515).

37. Zuo Y, et al. (2020) Prothrombotic autoantibodies in serum from patients hospitalized with COVID-19. Sci Transl Med 12(570).

38. Wang EY, et al. (2020) Diverse Functional Autoantibodies in Patients with COVID-19. medRxiv.

39. Hazenbos WL, et al. (1996) Impaired IgG-dependent anaphylaxis and Arthus reaction in Fc gamma RIII (CD16) deficient mice. Immunity 5(2):181–188.

40. Miyajima I, et al. (1997) Systemic anaphylaxis in the mouse can be mediated largely through IgG1 and Fc gammaRIII. Assessment of the cardiopulmonary changes, mast cell degranulation, and death associated with active or IgE- or IgG1-dependent passive anaphylaxis. J Clin Invest 99(5):901–914.

41. Syenina A, Jagaraj CJ, Aman SA, Sridharan A, & St John AL (2015) Dengue vascular leakage is augmented by mast cell degranulation mediated by immunoglobulin Fcgamma receptors. Elife 4.

42. Hallgren J, et al. (2007) Pulmonary CXCR2 regulates VCAM-1 and antigen-induced recruitment of mast cell progenitors. Proc Natl Acad Sci U S A 104(51):20478–20483.

43. Galdiero MR, et al. (2017) Bidirectional Mast Cell-Eosinophil Interactions in Inflammatory Disorders and Cancer. Front Med (Lausanne) 4:103.

44. Modin D, et al. (2020) Acute COVID-19 and the Incidence of Ischemic Stroke and Acute Myocardial Infarction. Circulation 142(21):2080–2082.

45. Li M, et al. (2004) Involvement of chymase-mediated angiotensin II generation in blood pressure regulation. J Clin Invest 114(1):112–120.

46. Miyazaki M, Takai S, Jin D, & Muramatsu M (2006) Pathological roles of angiotensin II produced by mast cell chymase and the effects of chymase inhibition in animal models. Pharmacol Ther 112(3):668–676.

47. Fyhrquist F & Saijonmaa O (2008) Renin-angiotensin system revisited. J Intern Med 264(3):224–236.

48. Ihara M, et al. (1999) Increased chymase-dependent angiotensin II formation in human atherosclerotic aorta. Hypertension 33(6):1399–1405.

49. Hamada H, et al. (1999) Increased expression of mast cell chymase in the lungs of patients with congenital heart disease associated with early pulmonary vascular disease. Am J Respir Crit Care Med 160(4):1303–1308.

50. Liu Y, et al. (2020) Clinical and biochemical indexes from 2019-nCoV infected patients linked to viral loads and lung injury. Sci China Life Sci 63(3):364–374.

51. Otani A, Takagi H, Oh H, Koyama S, & Honda Y (2001) Angiotensin II induces expression of the Tie2 receptor ligand, angiopoietin-2, in bovine retinal endothelial cells. Diabetes 50(4):867–875.

52. Parikh SM (2016) Targeting Tie2 and the host vascular response in sepsis. Sci Transl Med 8(335):335fs339.

