## Supplemental Materials for "Signatures of mast cell activation are associated with severe COVID-19"

[1]Program in Emerging Infectious Diseases, Duke-NUS Medical School, Singapore.
[2]Department of Pathology, Duke University Medical Center, Durham, NC, USA
[3]Duke-NUS Medical School, Program in Cardiovascular and Metabolic Disorders, Singapore
[4]Department of Infectious Diseases, Singapore General Hospital, Singapore
[5]Institute of Molecular and Cell Biology, A*STAR, Singapore
[6]Department of Medicine, Yong Loo Lin School of Medicine, National University of Singapore, Singapore
[7]Division of Infectious Disease, University Medicine Cluster, National University Hospital, Singapore
[8]Center for Applied Genomics and Precision Medicine, Duke University Medical Center, Durham, NC, USA.
[9]Division of Infectious Diseases, Duke University Medical Center; Durham VA Medical Center, Durham, NC, USA.
[10]Department of Anesthesiology, Duke University Medical Center
[11]Department of Microbiology and Immunology, National University of Singapore, Singapore
[12]SingHealth Duke-NUS Global Health Institute, Singapore

**This PDF file includes:**

Materials and Methods
Figs. S1 to S4
Table S1

### Materials and Methods

*SARS-CoV-2 Propagation*

Vero-E6 cells (ATCC®CRL-1586TM, 7.5x105 cells/mL) were infected with SARS-CoV-2 isolate hCoV-19/Singapore/2/2020 (WX-56) (GISAID accession ID: EPI_ISL_407987) at a multiplicity of infection (MOI) of 0.01 MOI for 1h at 37°C, 5% $CO_2$. After infection, 8 mL of DMEM medium with 5% fetal bovine serum (FBS) was added. Supernatant was harvested 3 days post infection. For plaque assays, Vero-E6 were seeded into 6-well plates ($6x10^5$ cells/well) and incubated overnight at 37°C, 5% $CO_2$ to achieve 90-100% confluence. SARS-CoV-2 isolate WX-56 was diluted up to $10^{-6}$ with 2% FBS DMEM medium. Cells were washed with 1mL PBS before infecting with 100 µL of diluted virus. The plate was incubated at 37°C, 5% $CO_2$ for 1h and rotated every 15 min to prevent the cells from drying. After incubation, 3 mL of carboxymethylcellulose (CMC) was added to each well and incubated further for 3 days at 37°C, 5% $CO_2$. The plate was fixed using 4% paraformaldehyde and stained with crystal violet 3 days post infection. To quantify virus using a median tissue culture infectious dose-50 ($TCID_{50}$) assay, Vero-E6 ($2x10^4$ cells/mL) were seeded into 96-well plates ($1x10^4$ cells/well) in quadruplets and incubated overnight at 37°C, 5% $CO_2$ to achieve 90-100% confluence. SARS-CoV-2 isolate WX-56 was diluted up to $10^{-8}$ with 5% FBS DMEM medium. Serially diluted virus (100 µL) was added to each well and incubated for 4 days before wells showing a cytopathic effect (CPE) were counted to determine the virus titer.

*Adeno-associated virus (AAV) production, purification and administration*

Viruses were produced as per standard protocol (Chew et al., Nature Methods, 2016). Briefly, AAVs were packaged via triple transfection of HEK293 cells. HEK293 cells were seeded in 10%FBS/DMEM supplemented with glutaMax (ThermoFisher Scientific #35050061), pyruvate, (ThermoFisher Scientific # 11360070) and MEM non-essential amino acids (Gibco # 11140050). Confluency at transfection was between 70–90%. Media was replaced with fresh pre-warmed growth media before transfection. For each HYPERFlask 'M' (Corning #CLS10034), 300 µg of pHelper (Cell Biolabs), 150 µg of pRepCap (encoding AAV9 (UPenn Vector Core)), and 150 µg of pAAV (containing the ITR-cargo-ITR) were mixed in 7 ml of DMEM, followed by mixing with 2.8 mg of PEI "MAX" 40k (Polysciences # 24765-1). The mixture was incubated at room temperature for 15 min, and transferred drop wise to the cell media. The day after transfection, the media was changed to DMEM containing 2%FBS, glutaMax, pyruvate, and MEM non-essential amino acids. Cells were harvested 48-72 hrs after transfection by dissociation with 5 mM EDTA in PBS (pH7.2), and pelleted at 1500 x *g* for 12 min. Cell pellets were resuspended in 5 ml of lysis buffer (Tris HCl pH 7.5, 2 mM $MgCl_2$,150 mM NaCl), and freeze-thawed three times between a dry ice-ethanol bath and a 37°C water bath. Cell debris was clarified by centrifuging 4000 x *g* for 5 min, and the supernatant collected. The collected supernatant was treated with 50 U/ml of Benzonase (Sigma-Aldrich) and 1 U/ml of RNase cocktail (Invitrogen # 10638255) for 30 min at 37°C to remove unpackaged nucleic acids. After incubation, the lysate was loaded on a discontinuous density gradient consisting of 4 mL, 6 mL, 7 mL, and 4 mL of 15%, 25%, 40%, and 60% Optiprep (Sigma-Aldrich # D1556) respectively in a 29.9 mL Optiseal polypropylene tube (Beckman-Coulter # 361625). The tubes were ultracentrifuged at 54,000 rpm, at 18 °C, for 1.5 h, on a Type 70 Ti rotor. The 40% fraction was extracted, and dialyzed with 0.001% pluronic acid/PBS, using Amicon Ultra-15 (100 kDa MWCO)(Millipore # UFC910024). The titre of the purified AAV9-hACE2 vector stocks were determined using real-time qPCR with ITR-sequence-specific primers and probe(*1*), referenced against the ATCC reference standard material 8 (ATCC).

C57Bl/6 mice that were treated with AAV9-hACE2 were purchased from InVivos, Singapore, and housed in the Duke-NUS Vivarium prior to use. Mice were anesthezized with xylazine/ketamine and inoculated intranasally with $0.5 \times 10^{11}$ PFU of AA9-ACE2 delivered in 30ul, alternating droplets between both both nares. SARS-CoV-2 infections were performed 21 days later to allow maximal expression of hACE2.

*Infection of AAV-hACE2 knock-in mice with SARS-Cov-*2

AAV-hACE2 knock-in mice were transferred to the Duke-NUS ABSL3 facility for SARS-CoV-2 infections. Mice were anesthetized with isoflurane for nasal inoculations. They were infected with $6x10^5$ $TCID_{50}$/mL of SARS-CoV-2 isolate WX-56 via nasal inoculation (6uL per nostril) and were subsequently weighed daily. Blood was collected on 1, 3, 5 and 7-days post-infection via cheek bleed. Mice were euthanized at 5 and 7 days post-infection and organs were harvested for RNA isolation and tissue sectioning. To isolate mouse serum, blood was allowed to clot at room temperature for 30 min prior to clarifying by centrifugation at 15000 rpm in a table top centrifuge. Mouse serum was inactivated by 30 minutes incubation at 56℃ to remove from the containment facility prior to further testing.

*Detection of MCPT1 in mouse serum*
Because mouse serum had been heat inactivated, potentially denaturing proteins, we used a western blot to detect MCPT1 levels in the blood. Serum was diluted 1:10 in PBS and denatured in 2x laemmli buffer (Bio-Rad, #1610737) before serum proteins were fractionated by SDS-PAGE. Proteins were then transferred on to PVDF membrane electrophoretically, which was blocked with 5% milk in TBST. Serum chymase was detected using Anti-Mast Cell Chymase antibody (Abcam, # ab2377,1:250) and Goat anti-Mouse IgG (H+L) Cross-Adsorbed Secondary Antibody, HRP (ThermoFisher Scientific, #G21040, 1:10000). Densitometric analysis was done using Fiji (ImageJ, NIH).

*SARS-CoV-2 Infection of NHPs*
Cynomolgus macaques (*Macaca fascicularis*) were used for NHP studies. Cynomolgus macaques were purchased from the SingHealth colony and free of antibodies against CoV. Infection studies were performed under BSL3 containment in the Duke-NUS Medical School ABSL3 facility. Prior to infection, NHPs were implanted with temperature transponders (Star-Oddi, Iceland). Body temperature was monitored every 15 min using a surgically implanted temperature sensor and rectally whenever the animals were anesthetized. For infection and sampling, animals were sedated with an intramuscular injection of ketamine (10-15 mg/kg) and medetomidine (0.05 mg/kg). Weight was recorded and a physical inspection was performed. Following initial sedation, 5% isoflurane was applied to achieve deeper anesthesia. A laryngoscope was used to intubate using an endotracheal (ET) tube. A 3 ml disposable luer lock syringe with 100 µl $3x10^7$ $TCID_{50}$/mL of SARS-CoV-2 isolate WX-56 was attached to ET tube connector for intratracheal infection. After injecting the virus, the ET tube was flushed with 1 ml of PBS to clear any residual inoculum. Animals were extubated and IV atipamezole was given to partially reverse medetomidine and facilitate faster recovery of the animals.

Post-infection, animals were observed twice daily for activity and observation of clinical signs. At days 0, 1, 3 5, 7, 9, 14 and 21 post-infection, the animals were anesthetized and intubated using the same technique used for infection. Animals were weighed and blood samples were collected from the femoral vein in CPT tubes. Nasal, rectal, throat and eye swabs were collected. Nasal rinse and lung lavage were performed with 500 µl and 6 ml PBS, respectively. The NHPs were euthanized at 21 days post-infection to allow for a full necropsy. Gross tissue observations were characterized by veterinarians and recorded upon necropsy. Blood, CSF, and tissues were harvested for RNA detection and histology.

*RNA isolation from mouse and primate tissues and PCR-based quantification of SARS-CoV-2*
Organs harvested from AAV-hACE2 knocked-in mice and NHPs were transferred to Lysing Matrix Y tubes (MPBio, #116960050-CF) containing 0.5 mm diameter Yttria-Stabilized Zirconium Oxide beads. 500 µL 5% FBS DMEM was added into each tube and tissues were homogenized with a handheld homogenizer (MPBio SuperFastPrep-1) for 1 min. Total RNA was extracted from all samples using E.Z.N.A. Total RNA Kit I (Omega Bio-tek) according to the manufacturer's instructions and samples were analysed by real-time quantitative reverse transcription-PCR (RT-qPCR) for the detection of SARS-CoV-2 in mouse and NHP samples as previously described(*2, 3*).

*Serum neutralizing antibody measurement in NHP by competitive ELISA*.
The cPass™ SARS-CoV-2 Surrogate Virus Neutralization Test Kit (GenScript) was used according to manufacturer's instructions. Briefly, each serum sample was diluted 1:10 in Sample Dilution Buffer and incubated with an equal volume of HRP-RBD solution for 30 min at 37°C. The mix was then applied to strips pre-coated with ACE2 protein for 15 min at 37°C. RBD-ACE2 binding was visualized by addition of TMB substrate for 15 min at room temperature. The reaction was terminated using Stop Solution and absorbance measured at 450 nm. Inhibition of RBD-ACE2 binding was calculated using the formula: (1-(OD value of sample)/(OD value of negative control))x 100%.

*Histology*
Paraformaldehyde fixed tissues were snap frozen in O.C.T compound (Tissue-Tek, Sakura) and sectioned to 15 µm thickness. For the toluidine blue staining protocol to identify MCs, sections were fixed

in Carnoy's solution for 30 min at room temperature. Following fixation, sections were stained using 0.1% toluidine blue stain (Sigma-Aldrich, #198161) for 20 min and excess dye was removed by gently washing in running tap water followed by rinsing in distilled water. Sections were then dehydrated quickly in 95% alcohol followed by 2 changes in 100% alcohol and mounted using permanent mounting medium (VectaMount, #H-5000). For hematoxylin and eosin staining, air dried sections were rehydrated using a graded series of alcohol and stained using modified Harris hematoxylin (Sigma-Aldrich, #HHS32) for 10 min followed by a wash in tap water and two changes in distilled water. Sections were briefly dipped in 1% acid alcohol solution and quickly rinsed in distilled water before differentiating using 0.05% lithium carbonate solution for 1 minute. Sections were washed in distilled water and dehydrated using 95% alcohol followed by a counter stain using 0.25% eosin y (Sigma-Aldrich, #HT110232). Finally, sections were rinsed in 95% alcohol to remove excess eosin stain followed by 2 changes in 100% alcohol and air dried before mounted using permanent mounting medium (VeactaMount, #H-5000). Images were obtained using a light microscope (Nikon) and processed using ImageJ Fiji.

*Immunostaining of MCs*
Tissue sections were permeabilized using 0.3% Triton X-100 in PBS for 30 min at room temperature followed by incubation with blocking buffer (0.1% Saponin+ 5% BSA in PBS) for 2h at room temperature. Mast cells were probed using heparin binding Avidin conjugated to FITC (BD Pharmingen, #554057) for overnight at 4°C. Sections were washed 3-4 times using PBS before mounting using Fluoroshield mounting medium containing DAPI (Sigma-Aldrich, #F6057). Images were acquired using THUNDER Imaging Systems (Leica).

*Clinical samples and microarray analysis*
The data associated with human transcriptional responses was approved by the SingHealth Combined Institutional Review Board (CIRB 2017/2374). The detailed study design and protocol has been described previously(*4*), where whole blood transcript expression was measured in the severe and mild COVID-19 patients by the Affymetrix GeneChip Human Gene 2.0 ST Array. The raw data for the microarray profiling is available at Array Express (E-MTAB-9721), and the log2 counts are generated by the Transcriptome Analysis Console (Thermo Fisher), analyzed between the different days relative to peak severity with regards to respiratory function (Day 0). Temporal gene expression was analyzed by EDGE based on the log2 intensity counts(*5*), and genes that were significantly altered in the severe COVID-19 patients were identified based on p-value and q-value < 0.05. The genes from the MC-specific and the MC/Basophil phenotype were obtained from(*6*), and Partek® Genomics Suite® was used to tabulate the Least Square Means (LSMeans) values. Genes of increased expression during the acute phase or recovery phase were then further stratified. Normalized expression was tabulated by taking the average LSMeans values of all MC and MC/Basophil phenotype genes that were of increased expression during the acute phase. Heatmaps and graphs were constructed using Prism 9.0.2 software.

For analysis of the microarray or nCounter datasets, *Z*-score transformation was performed as described previously[43]. To identify DEGs between symptomatic and asymptomatic subjects at baseline, Partek Genomics Suite Analysis v.7 software was used and Bonferroni's correction was performed based on the total number of 34,667 genes that were detected by microarray, using $P < 0.05$. No cutoff on fold change was imposed. For pathway analysis, the identified DEGs were used as input data, and analyzed against the Reactome database using the Enrichr tool[17]. Both *P* values and combined scores for each enriched pathway were obtained from the Enrichr tool analysis using algorithms that are described in greater detail by Kuleshov et al.[17]. Volcano plots were constructed using Prism v.8.1.0 software. To evaluate whether there was any statistically significant difference in specific Reactome pathways between symptomatic and asymptomatic subjects, the average *Z*-scores of all genes in each of the pathway were plotted. An unpaired, Student's *t*-test was then used to assess the statistical significance of the observed differences. The ROC curves for the various pathways were also determined using the average *Z*-scores of all genes in the UPR, sumoylation and TCA cycle pathway and plotted using Prism v.8.1.0. Ingenuity software was used to generate gene network diagrams.

*Clinical samples for ELISA analyses*

Patients with confirmed SARS-CoV-2 infection in either the hospital or outpatient setting were identified through the Duke University Health System (DUHS) or the Durham Veterans Affairs Health System (DVAHS) and enrolled into the Molecular and Epidemiological Study of Suspected Infection (MESSI, Pro00100241). Written informed consent was obtained from all subjects or legally authorized representatives. All plasma samples were from the time of enrollment with the exception of 1 patient whose sample was from day 3 and two subjects whose samples were from day 7 after enrollment. Peripheral blood was drawn into EDTA vacutainers, centrifuged, and isolated plasma was stored at −80°C until analysis. COVID-19 samples were processed under BSL2 with aerosol management enhancement and the following ELISAs were performed: chymase (Antibody Online, Aachen, Germany), Angiopoietin-1 and- 2 (ThermoFisher Science, Waltham, MA).

Patients with confirmed SARS-CoV-2 infection in Singapore were recruited in accordance with protocols approved by the intuitional IRB, DSRB domain E, (#2020/00120) and informed consent was taken from all patients. Serum samples from acute patients, <7 days of illness, were tested for chymase using the Human mast cell chymase I (CMA-I) kit (BlueGene Biotech, catalogue number E01M0368), according to manufacturer's instructions. Chymase concentration values obtained and previously published using the same kit for healthy control and DENV patients from Singapore(7) were compared to the values obtained in SARS-CoV-2 patients.

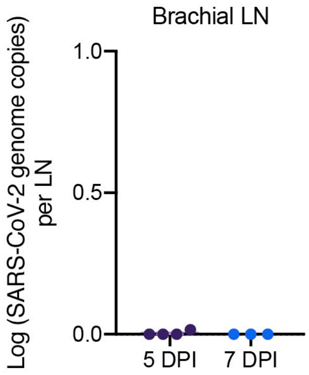


**Supplemental Figure 1. No detection of SARS-CoV-2 in mouse brachial LNs days 5 or 7 post-infection**

A

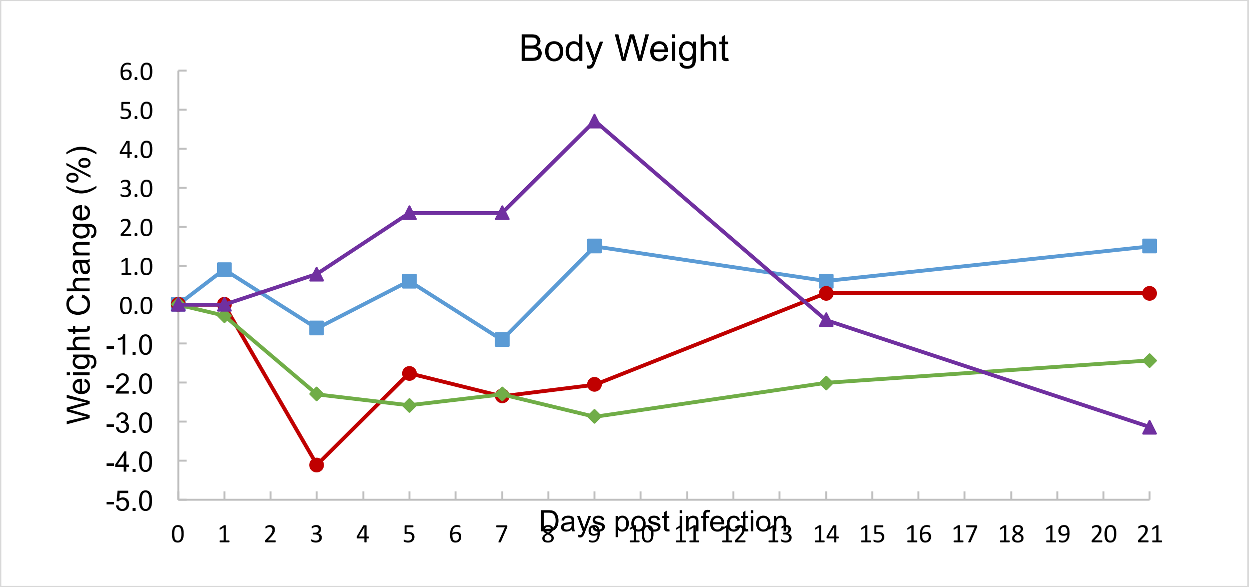


B

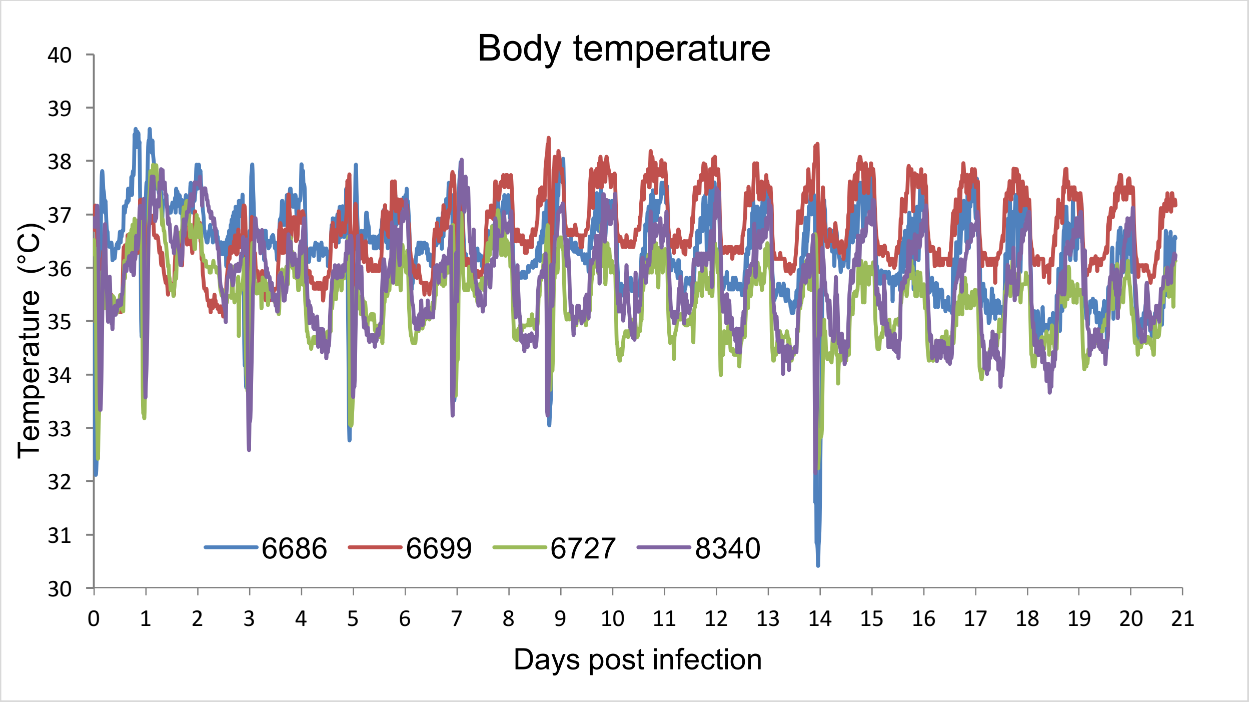


**Supplemental Figure 2: Body weight and temperature in cynomolgus macaques infected with SARS-CoV-2.** A) Changes in body weight (% of initial weight) after infection with SARS-CoV-2. B) Body temperature measured every 15 min by telemetry after infection. Dips in body temperature on D0, 1, 3, 5, 7, 9, and 14 are due to anesthesia administered for sample collection.

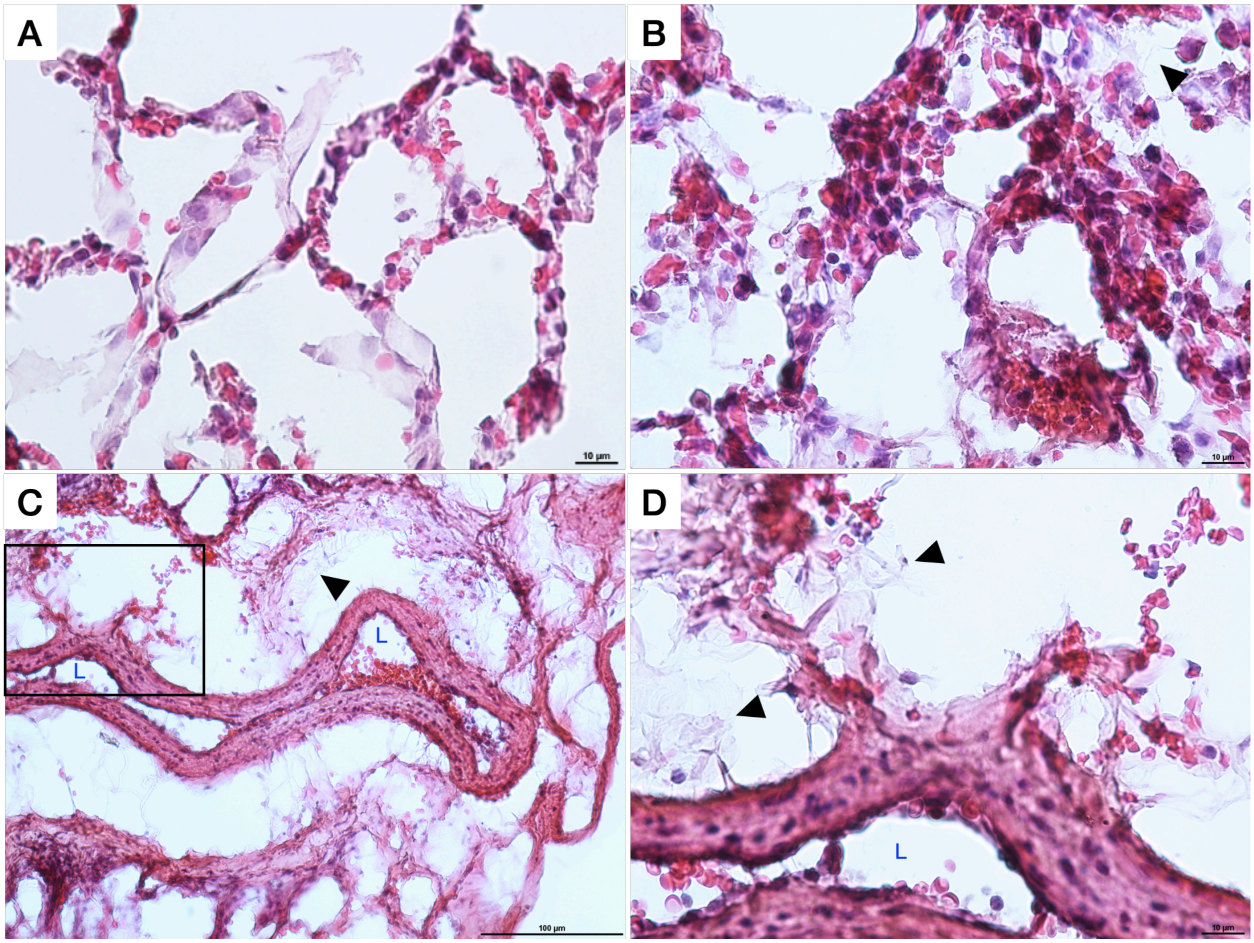


**Supplemental Figure 3: Observation of hemorrhaging and coagulation in SARS-CoV-2-infected NHP lung alveoli.** (**A-B**) Representative images showing the presence of free RBCs in the alveoli of NHPs 21d after SARS-CoV-2 infection. Notable thickening of the alveoli walls is observed in some sections, as in panel **B**. (**C**) Hemorrhaging from an artery in the lung is shown at low magnification (10x) and reimaged at higher magnification in panel **D**, corresponding to the boxed inset in **C**. For **B-D**, black arrows indicate examples of fibrin deposition in the tissue and for **C-D**, the artery lumen is indicated by a blue “L”.

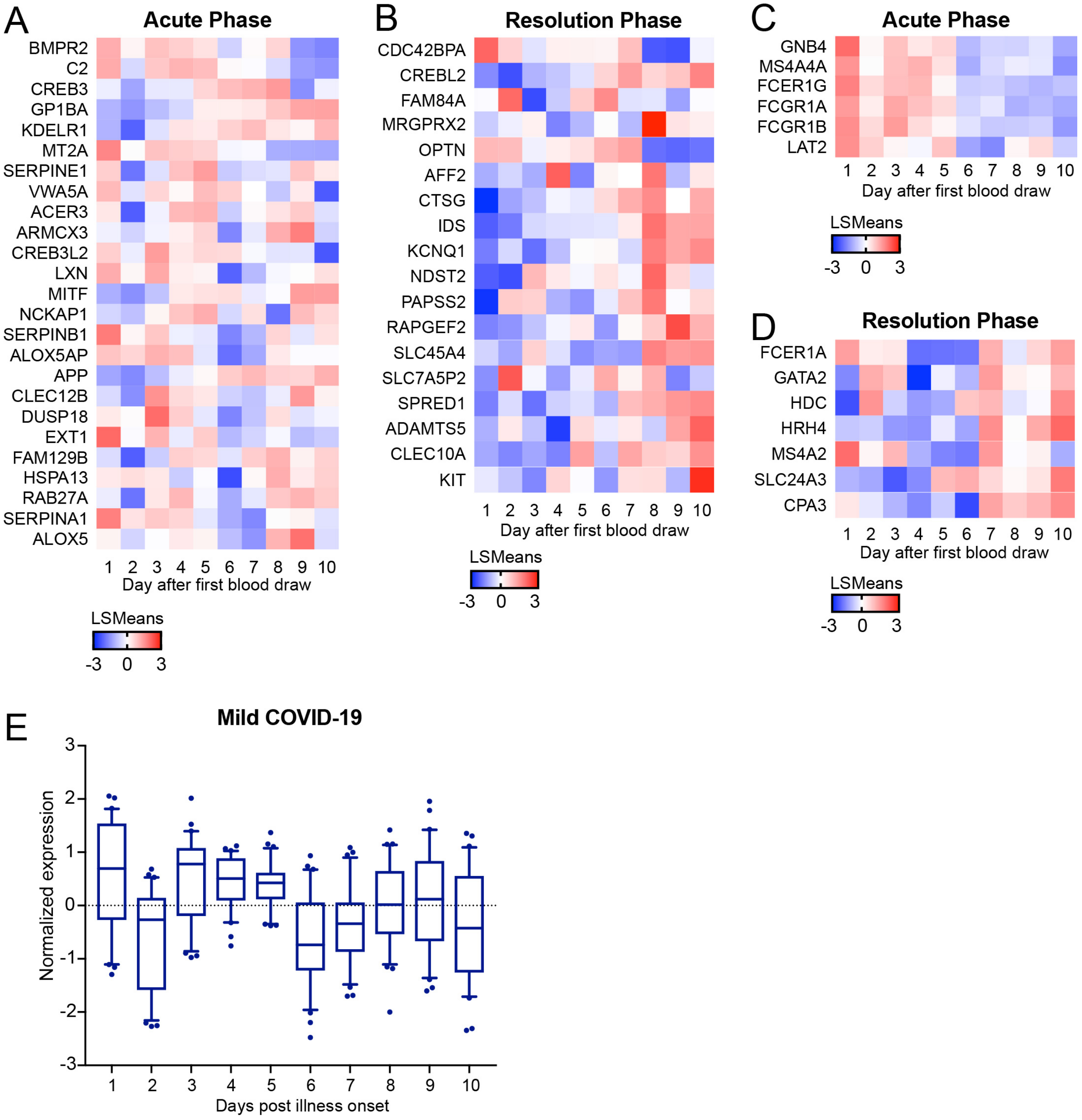


**Supplemental Figure 4: Transcriptional responses of MC-associated genes in mild COVID-19 patients.** Genes associated with a (**A-B**) MC-specific or (**C-D**) MC/basophil phenotype that were significantly regulated in severe COVID-19 patients are presented by heat map throughout the course of mild COVID-19 disease. Heatmap shows the LSmean expression values of mild COVID-19 patients (n=3) over the course of illness. (**E**) Normalized expression levels of MC-specific expression in mild COVID-19 patients through the course of disease.

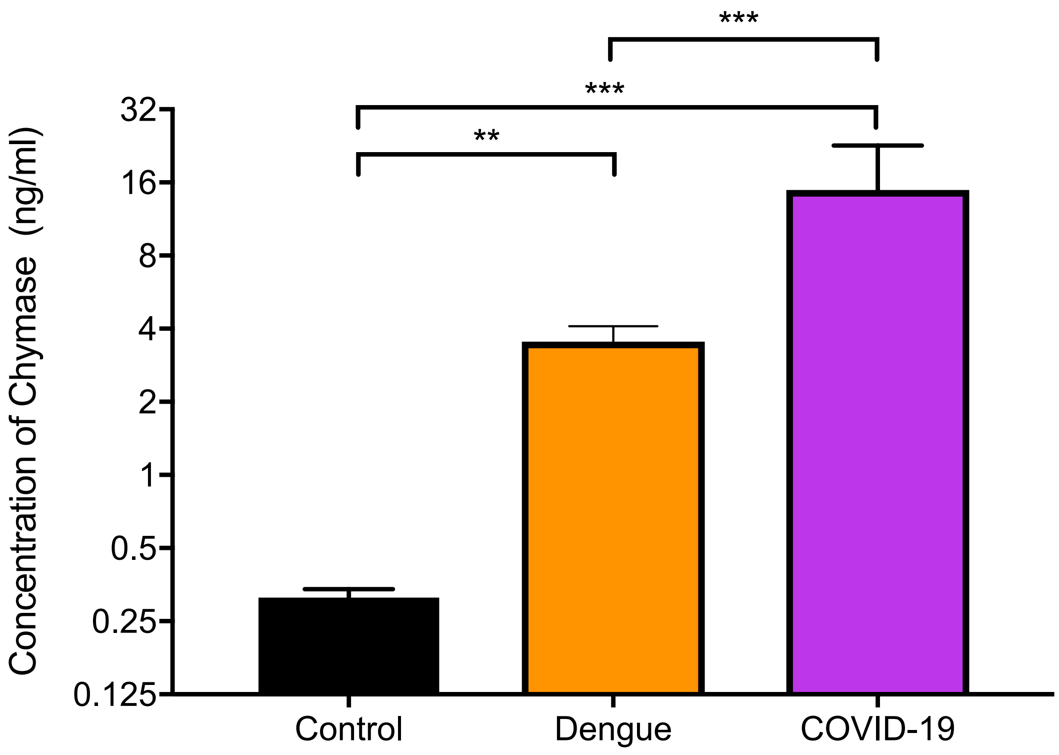


**Supplemental Figure 5: Chymase detection in COVID-19 patients.** Levels of chymase in the serum of acute COVID-19 patients recruited in Singapore were compared to the concentrations previously detected and reported in a study of acute dengue patients(7), and to healthy controls. Concentrations were compared by 1-way ANOVA with Bonferroni's post-test to determine p-values. N=10 for controls, N=108 for dengue and N=3 for COVID-19. For **, $p<0.01$; for ***, $p<0.001$.

| | % Neutralization | | | |
|---|---|---|---|---|
| DPI | **NHP 8340** | **NHP 6686** | **NHP 6699** | **NHP 6727** |
| 0 | 0.00 | 1.31 | 8.10 | 19.64 |
| 1 | 0.00 | 3.71 | 8.93 | 17.87 |
| 3 | 0.00 | 6.26 | 8.47 | 19.00 |
| 5 | 4.88 | 6.15 | 17.71 | 17.64 |
| 7 | 0.97 | 3.75 | 19.85 | **20.33*** |
| 9 | 5.23 | **38.75*** | **38.87*** | **47.64*** |
| 14 | **75.22*** | **89.75*** | **73.24*** | **86.37*** |
| 21 | **85.68*** | **91.37*** | **71.24*** | **88.76*** |

**Table S1: Surrogate virus neutralization test.** Neutralizing activity was determined using an ELISA-based cPass™ kit that assessed antibodies blocking the interaction between RBD and ACE2 receptor. A cut-off of 20% inhibition (*) is used to identify seropositive samples.


1. C. Aurnhammer *et al.*, Universal real-time PCR for the detection and quantification of adeno-associated virus serotype 2-derived inverted terminal repeat sequences. *Hum Gene Ther Methods* **23**, 18-28 (2012).
2. V. M. Corman *et al.*, Detection of 2019 novel coronavirus (2019-nCoV) by real-time RT-PCR. *Euro Surveill* **25**, (2020).
3. X. Lu *et al.*, US CDC Real-Time Reverse Transcription PCR Panel for Detection of Severe Acute Respiratory Syndrome Coronavirus 2. *Emerg Infect Dis* **26**, (2020).
4. E. Z. Ong, Kalimuddin, S., Chia, W.C., Ooi, S.H., Koh, C.W.T., Tan, H.C., Zhang, S.L., Low, J.G., Ooi, E.E., Chan, K.R., Temporal dynamics of the host molecular responses underlying severe COVID-19 progression and disease resolution. *Ebiomedicine*, (in press).
5. J. D. Storey, W. Xiao, J. T. Leek, R. G. Tompkins, R. W. Davis, Significance analysis of time course microarray experiments. *Proc Natl Acad Sci U S A* **102**, 12837-12842 (2005).
6. D. F. Dwyer, N. A. Barrett, K. F. Austen, C. Immunological Genome Project, Expression profiling of constitutive mast cells reveals a unique identity within the immune system. *Nat Immunol* **17**, 878-887 (2016).
7. A. L. St John, Rathore, A. P. S., Raghavan, B., Ng, M. L., Abraham, S. N., Contributions of mast cells and vasoactive products, leukotrienes and chymase, to dengue virus-induced vascular leakage. *eLife*, (2013).
